# Reconciling epidemiological models with misclassified case-counts for SARS-CoV-2 with seroprevalence surveys: A case study in Delhi, India

**DOI:** 10.1101/2020.07.31.20166249

**Authors:** Rupam Bhattacharyya, Ritwik Bhaduri, Ritoban Kundu, Maxwell Salvatore, Bhramar Mukherjee

**Affiliations:** Department of Biostatistics, University of Michigan, Ann Arbor, MI 48109, USA; Indian Statistical Institute, Kolkata 700108, West Bengal, India; Center for Precision Health Data Science, University of Michigan, Ann Arbor, MI 48109, USA

## Abstract

Underreporting of COVID-19 cases and deaths is a hindrance to correctly modeling and monitoring the pandemic. This is primarily due to limited testing, lack of reporting infrastructure and a large number of asymptomatic infections. In addition, diagnostic tests (RT-PCR tests for detecting current infection) and serological antibody tests for IgG (to assess past infections) are imperfect. In particular, the diagnostic tests have a high false negative rate. Epidemiologic models with a latent compartment for unascertained infections like the Susceptible-Exposed-Infected-Removed (SEIR) models can provide predictions for unreported cases and deaths under certain assumptions. Typically, the number of unascertained cases is unobserved and thus we cannot validate these estimates for a real study except for simulation studies. Population-based seroprevalence studies can provide a rough estimate of the total number of infections and help us check epidemiologic model projections. In this paper, we develop a method to account for high false negative rates in RT-PCR in an extension to the classic SEIR model. We apply this method to Delhi, the national capital region of India, with a population of 19.8 million and a COVID-19 hotspot of the country, obtaining estimates of underreporting factor for cases at **34-53 times** and that for deaths at **8-13 times**. Based on a recently released serological survey for Delhi with an estimated 22.86% seroprevalence, we compute adjusted estimates of the true number of infections reported by the survey (after accounting for misclassification of the antibody test results) which is largely consistent with the model outputs, yielding an underreporting factor for cases from **30-42**. Together with the model and the serosurvey, this implies approximately **96-98% cases in Delhi remained unreported** and whereas only 109,140 cases were reported on July 10, the true number of infections varied somewhere between **4.4-4.6 million** across different estimates. While repeated serological monitoring is resource intensive, model-based adjustments, run with the most up to date data, can provide a viable option to keep track of the unreported cases and deaths and gauge the true extent of transmission of this insidious virus.

## INTRODUCTION

COVID-19 was first diagnosed in Wuhan, China in December 2019 and was quickly declared a pandemic by the World Health Organization on March 11^1^. The first case in India was declared on January 30, and as of July 28, there have been 1,531,783 cases and 34,224 deaths reported^2^. India responded quickly, instituting a nationwide lockdown on March 25, when there were only 657 cases and 11 deaths^2-3^. However, given the high rate of asymptomatic individuals and limited and imperfect diagnostic testing, it is unclear how many people have actually been infected.

Classical epidemiologic models, like a susceptible – exposed – infected – removed (SEIR) compartmental model, have been used to predict the growth of the COVID-19 pandemic. For example, a modification of the standard SEIR model – which accounts for pre-symptomatic infectiousness, time-varying ascertainment rates, transmission rates and population movements – applied on data from Wuhan, China, identified that the outbreak had high covertness and high transmissibility^4^. They estimated that 87% (with a lower bound of 53%) of the infections in Wuhan before March 8 were unascertained^4^. However, traditional models, including the one used in this paper, do not take into account the underreporting and potential misclassification via imperfect testing.

In the context of identification of cases, there are two classes of tests that are being discussed in the literature: diagnostic tests and antibody tests. A diagnostic test (typically an **RT-PCR test**) is used to identify the presence of SARS-CoV-2, indicating an *active* infection^5^. An antibody test, i.e., a **serology test**, looks for the presence of antibodies, the body’s immune response to fight off SARS-CoV-2, indicating a *past* infection^6^. Figure 1 exhibits a timeline in terms of when these tests are done during the time course of an infection. Due to a large number of asymptomatic cases and limited number of tests, many infections do not get detected. Population-based seroprevalence surveys, therefore, give us an idea about the “true number of infections” including reported and unreported cases, and consequently, the ascertainment rate^6^. Thus, adjusted estimates of total number of cases and ascertainment rates based on serological surveys when available provide an option to validate model-based estimates of unreported cases and underreporting factors, which would usually be impossible to validate (except for a simulation study) since these numbers are not observable the real data.

**Figure 1.**
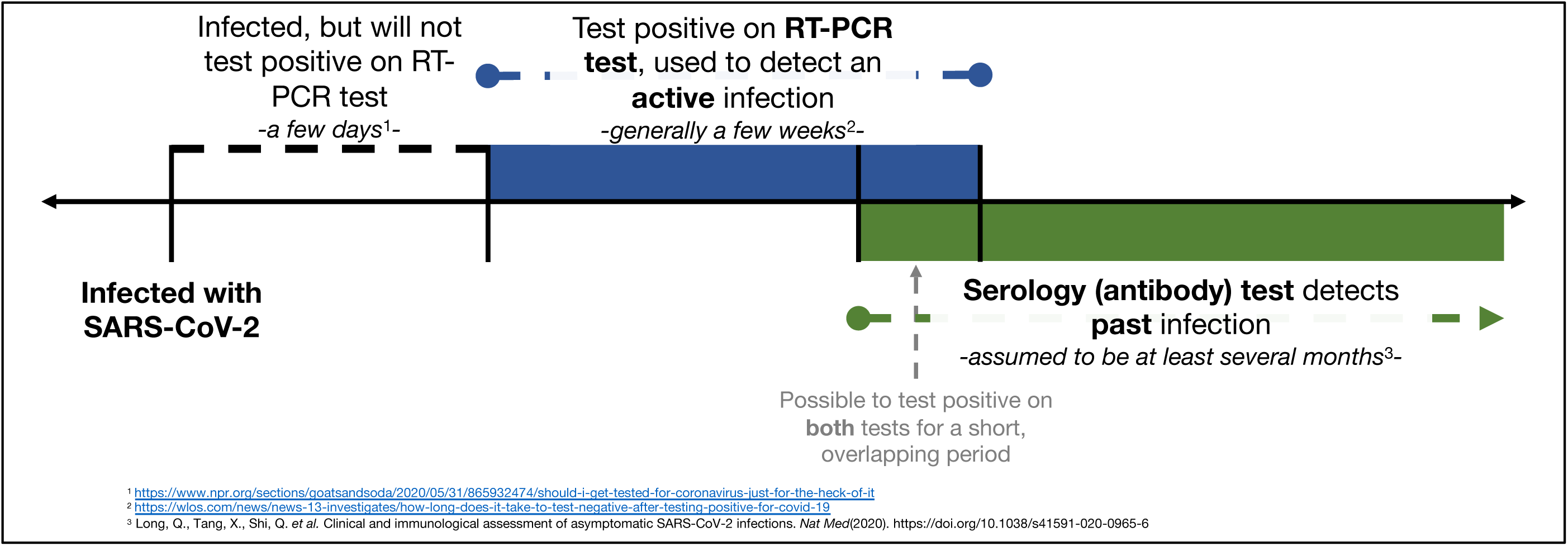
Timeline of COVID-19 diagnostic and antibody tests with respect to the infection and immune response time frame.

In an attempt to understand the spread of the virus in Delhi, the national capital region of India and one of the hotspots of COVID-19 in the country, the National Centre for Disease Control (NCDC) in India performed a serological survey in Delhi. While limited on reported details, the Delhi Serology Study collected 21,387 random samples across 11 districts in Delhi between June 27 and July 10 and found COVID-19 antibodies present in 22.86% of samples^7-9^. A simple proportional estimate would tell us that Delhi, with approximately 19.8 million people, had a total number of cases standing somewhere around 4.6 Million by July 10. This contrasts sharply both with the 109,140 cumulative cases (3,300 total deaths) reported in Delhi as of July 10, which represents roughly 0.55% of Delhi’s population, indicates that roughly, only 2.4% of cases are being detected (underreporting factor of about 42), and also implies that the infection fatality rate (IFR) for Delhi is of the order of 0.07% or 717 per million. This IFR seems low compared to estimates worldwide^10^ and as such it may be reasonable to argue that COVID related deaths are also possibly unreported, or the cause of death misclassified. The doubt regarding death data is further substantiated as a very small fraction of deaths in India are medically reported^11^ and the IFR estimates for SARS-CoV-2 from other studies in the world^10^ appear to be higher than influenza (infection fatality rate of influenza, as of 2018-19, is at 961 per million or around 0.1%)^12^

Both diagnostic and antibody tests suffer from the issue of false negatives and false positives, and depending on which test is being talked about, one or the other of these errors are more crucial. For the RT-PCR test, false negatives are more worrisome since that means allowing an infected person to go about freely, potentially spreading the virus around. Similarly, we worry about the false positives of an antibody test more, since it gives the false impression that the person has been infected in the past, gained immunity, and is unlikely to be infected again. The RT-PCR test is quoted to have a high false negative rate, ranging from 15-30% (i.e., low sensitivity, 85-70%), and a low false positive rate around 1-4% (i.e., high specificity, 99-96%)^13^. The antibody test assays are more precise - the commercial assays have sensitivity around 97.6% and specificity of 99.3% (DiaSorin) at about 15 days after infection^14^. The ELISA assay used in the Delhi serosurvey is a customized assay, about which no official information was not publicly released, but some discussions on imperfections of the test are available in public media domain^15^. In light of these imperfections and the high rate of asymptomatic COVID-19 cases, we develop an extension of a standard SEIR model incorporating misclassification due to imperfect diagnostic testing to predict both the number of unreported cases and deaths and to estimate the rate at which COVID-19 cases are being underreported. An alternative approach that has been discussed in contemporary literature is to model test activities in a way such that symptomatic individuals are identified and successfully isolated with a given average delay from the onset of symptoms^16^. This approach can handle both real-time RT-PCR tests and lab-based sero-surveillance.

Because the Delhi Serology Study provides a seroprevalence estimate, this is a unique opportunity to help validate predictions for latent unreported infections for a SEIR model. We perform adjustments of the reported case counts (and hence corresponding metrics such as the underreporting factor, infection-fatality rate and case-fatality rate) under different sensitivity and specificity assumptions for the diagnostic and antibody tests and potential underreporting of the death counts, and validate the model-based estimates of the extent of underreporting to those obtained from the seroprevalence-based calculations. We apply this framework to Delhi, using reported COVID-19 data from covid19india.org^2^. This framework can be adapted to and applied outside of Delhi and in other contexts where imperfect and limited testing exists.

## RESULTS

### Extended SEIR Model Adjusted for Misclassification

Under low (0.7), medium (0.85) and perfect (1) sensitivity, and perfect (1) specificity assumptions for the RT-PCR diagnostic test, we preform predictions of total (reported and unreported) cases and deaths for Delhi using the proposed extended SEIR model. Using data till June 30, this model estimates 4.8 million cases and 33,165 deaths on July 10 if we assume the RT-PCR test has a sensitivity of 0.85, and those predicted counts become 4.2 million and 28,499, respectively, if the sensitivity is assumed to be 1.0. Compare to the observed case and death counts of 109,140 and 3,300 reported in Delhi as of July 10.^2^ The model predictions under the different scenarios considered and the performances in terms of fitting the daily observed case and death counts are summarized in Figures 2-3. Looking at the ratio of predicted total number of cases and the predicted number of reported cases on July 10, it appears that the underreporting factor for cases reported by the model is within the range of **34-53** and the same for the deaths is between **8-13** (Table 1 and Figure 4). This implies according to the model **97-98%** Delhi’s cases remain undetected.

**Table 1.** Summary of extended SIR model results for Delhi. Predicted cumulative case and death counts and corresponding underreporting factors with respect to the observed data are presented for July 10. The specificity of the RT-PCR test is assumed to be 1. The observed number of cumulative cases and deaths in Delhi on July 10 were taken to be 109,140 and 3,300 respectively, according to covid19india.org.

| **Sensitivity**<br>**of RT-**<br>**PCR Test** | **Predicted**<br>**Reported**<br>**Cases** | **Predicted**<br>**Total**<br>**Cases** | **Under-**<br>**reporting**<br>**Factor for**<br>**Cases** | **Predicted**<br>**Reported**<br>**Deaths** | **Predicted**<br>**Total**<br>**Deaths** | **Under-**<br>**reporting**<br>**Factor for**<br>**Deaths** |
| --- | --- | --- | --- | --- | --- | --- |
| **0.7** | 119,920 | 6,318,663 | 52.6 | 3,386 | 43,978 | 12.9 |
| **0.85** | 119,879 | 4,780,982 | 39.8 | 3,384 | 33,165 | 9.8 |
| **1** | 119,603 | 4,164,568 | 34.8 | 3,376 | 28,499 | 8.4 |

**Figure 2.**
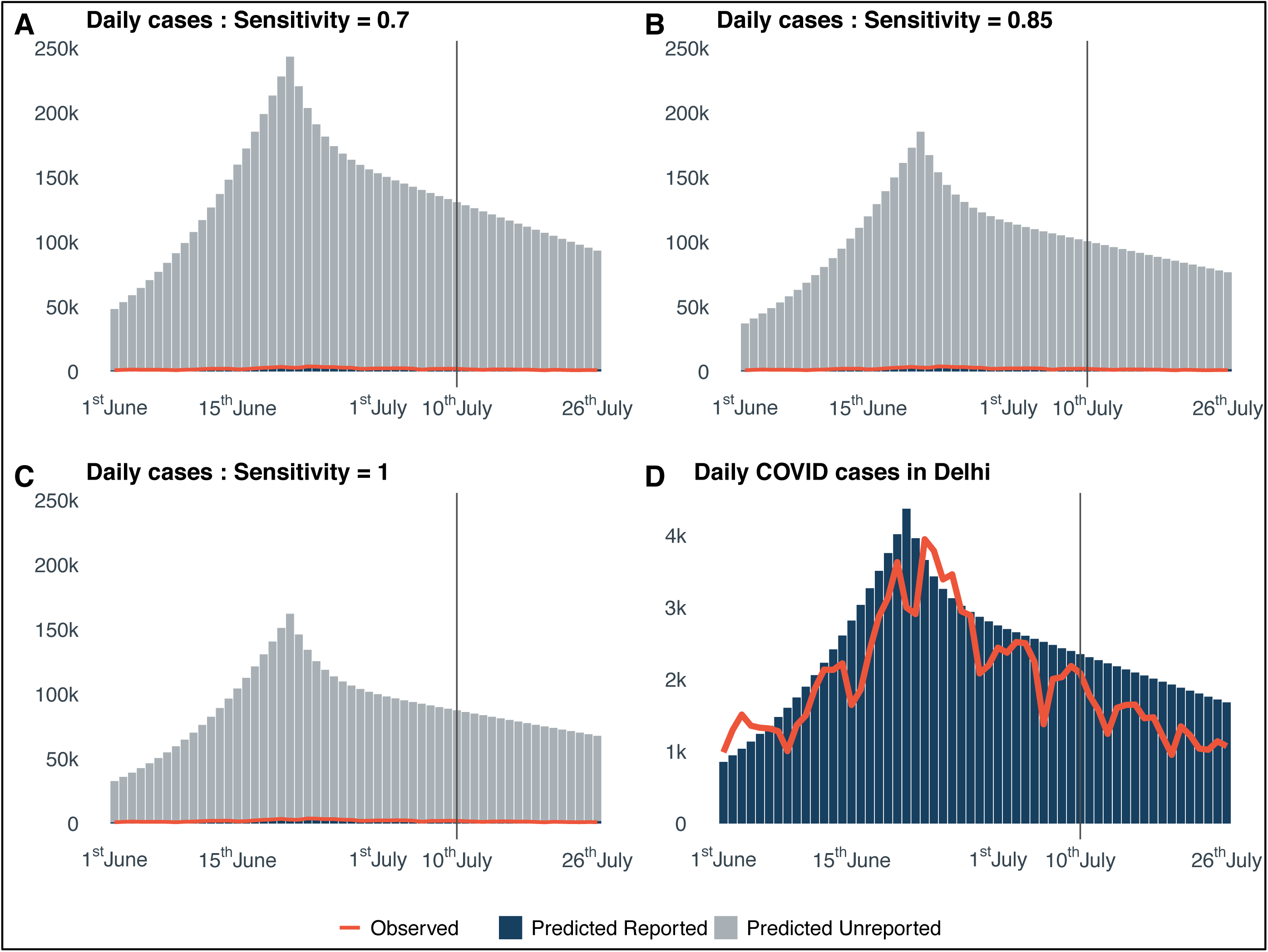
Bar plots of predicted reported and unreported daily cases from June 1 to July 26. Panels A, B and C depict the predictions under assumed sensitivity of the diagnostic test at 0.7, 0.85 and 1, respectively. Panel D shows the consistency of the predictions with the observed data. The specificity of the diagnostic test is assumed to be 1.

**Figure 3.**
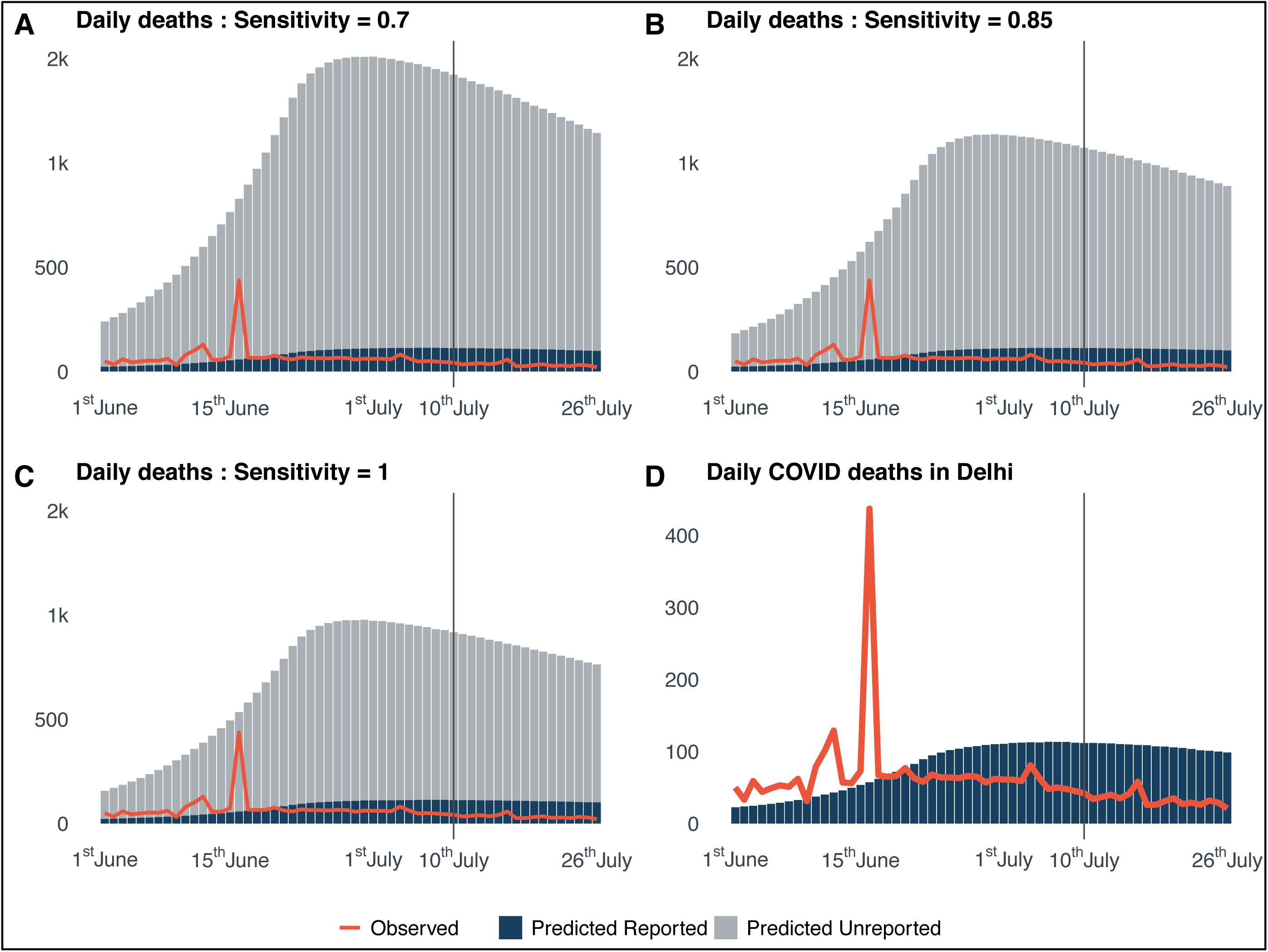
Bar plots of predicted reported and unreported daily deaths from June 1 to July 26. Panels A, B and C depict the predictions under assumed sensitivity of the diagnostic test at 0.7, 0.85 and 1, respectively. Panel D shows the consistency of the predictions with the observed data. The specificity of the diagnostic test is assumed to be 1.

**Figure 4.**
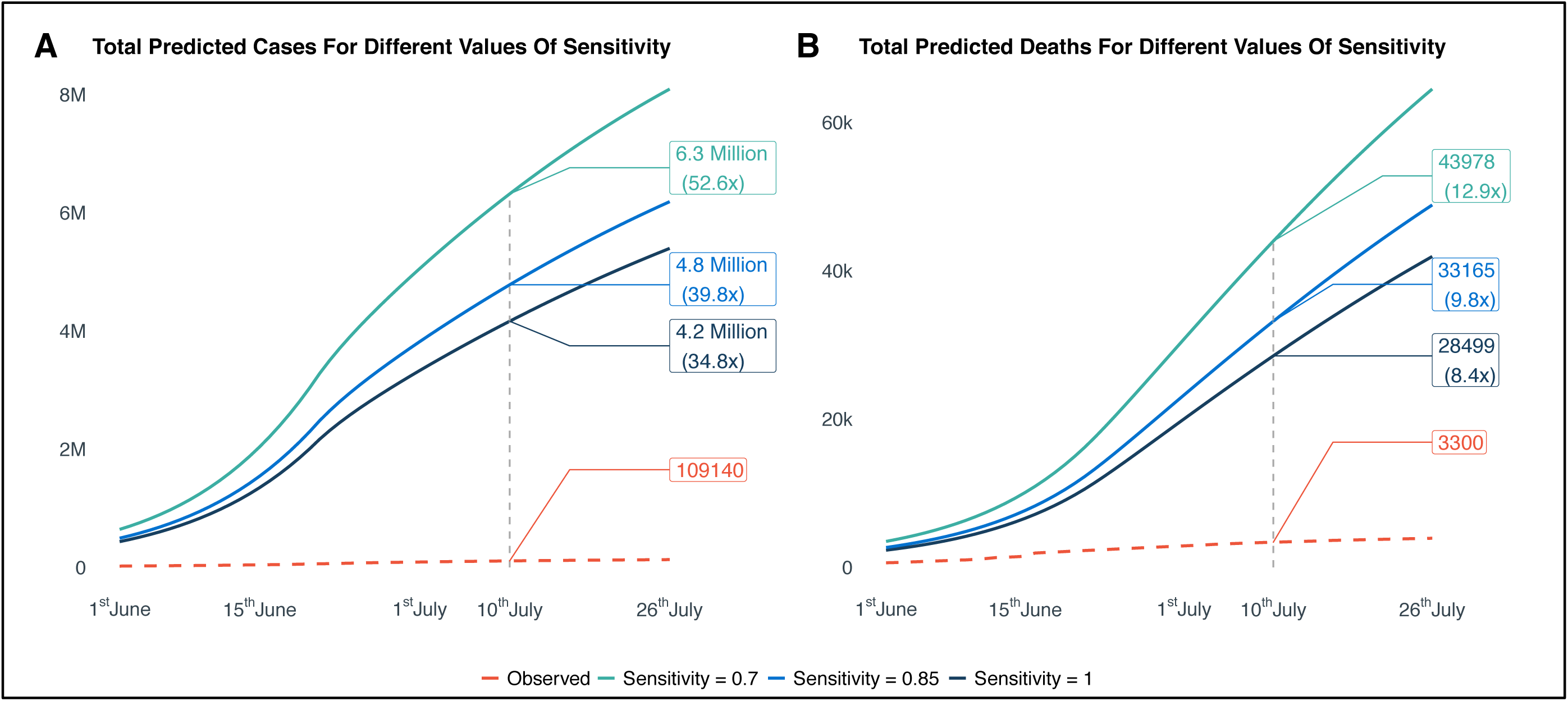
Summary of cumulative total (reported and unreported) cases and deaths for three different assumed values of specificity for the diagnostic test: 0.7, 0.85, 1. Panel A and B respectively summarize the cases and deaths, along with their reported observed counterparts. The specificity of the diagnostic test is assumed to be 1.

### Naïve Corrections to Reported Test Results using Known Misclassification Rates for Tests

Since the unreported number of cases and subsequently, the underreporting factor, are not part of the observed data and therefore cannot directly be validated, we validate these estimates using adjustments regarding the imperfection of the tests and estimated number of true infections predicted by the serosurvey data. We also consider the hypothetical scenario considering 10-fold underreporting of deaths, as suggested by the model outputs. However, we are not able to perform any validation for the estimated underreporting factor for deaths as we do not have estimate of true death rates or excess deaths.

Using varying (low to perfect) sensitivities and specificities for the diagnostic and antibody tests, we estimate that the true case count in Delhi as of July 10 lies between **4.4 and 4.6 million**, which represents **30 to 42 time**s the number of reported cases (Table 2), *these estimates being greatly in agreement with the model outputs, as reported in the previous subsection*. This indicates **96-97%** cases in Delhi were underreported.

**Table 2.**
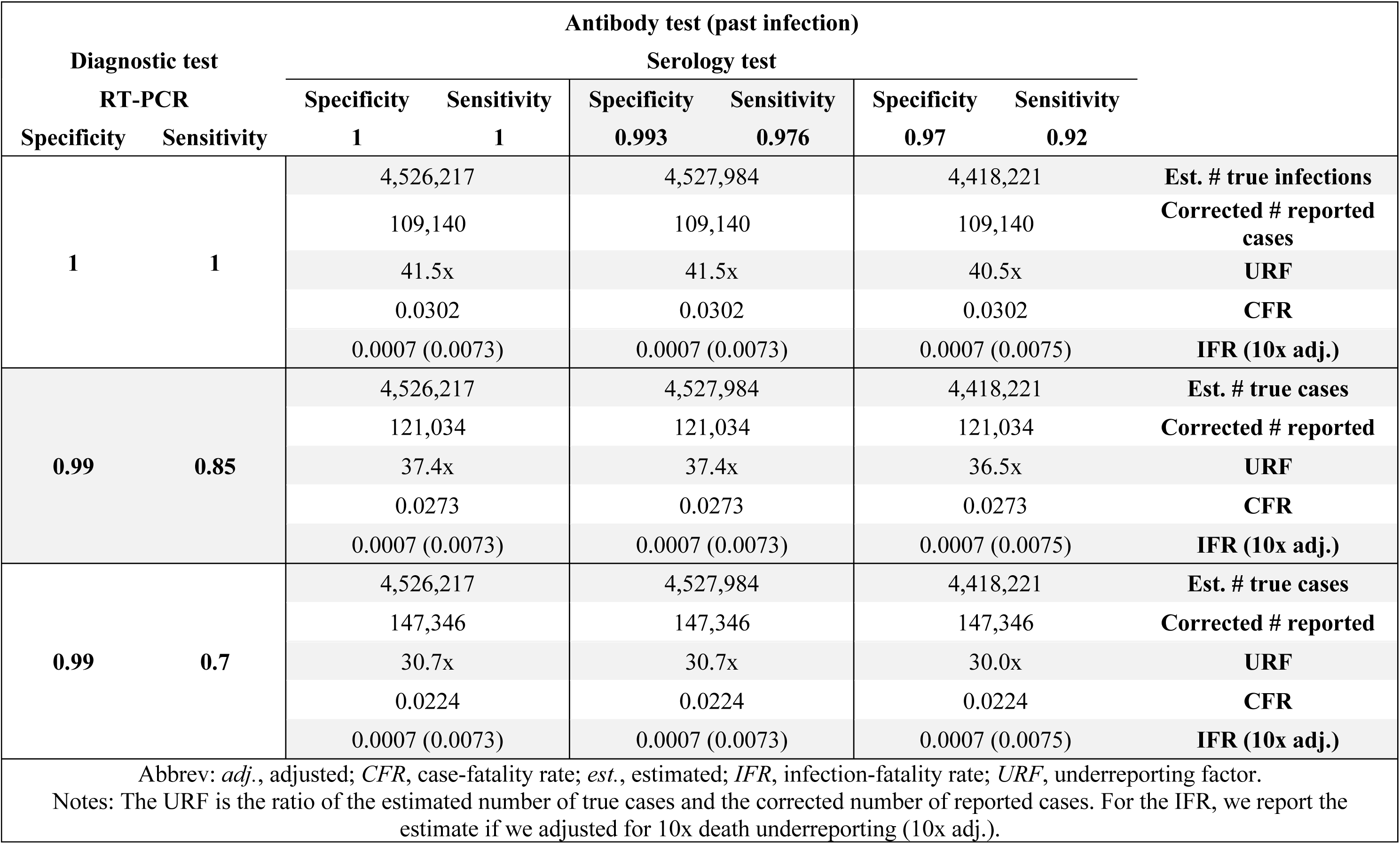
Summary of corrected number of cases, estimated underreporting factor, case-fatality rate based on reported cases and infection-fatality rate across different testing scenarios. Population size of Delhi is collected from https://censusindia.gov.in/, and the testing, infection, recovery and fatality data are taken from https://covid19india.org/.

### Case fatality rate (CFR) and Infection fatality rate (IFR)

The sensitivity and specificity of the diagnostic test impact our estimate of the case-fatality rate 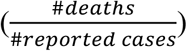, but not the infection-fatality rate 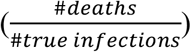. We estimate that the CFR lies between 2.24-3.02%. On the other hand, the sensitivity and specificity of the antibody test impact our estimates of the IFR. We estimate that the IFR lies between 0.07-0.08% based on the reported death counts. If we consider a 10-fold underreporting in death counts, the infection-fatality rate estimate increases to 0.7-0.8% (Table 2).

## DISCUSSION

We developed an extension of the standard SEIR compartmental model to adjust for the misclassifications due to imperfect diagnostic testing. Applying our model on publicly available infection and death data for Delhi, we estimated the underreporting factor for cases to be somewhere between **34 and 53** and that for deaths to be somewhere between **8 and 13** on July 10. Further, using adjustments under different imperfection scenarios for the diagnostic and antibody tests, we came up with adjusted estimates of the underreporting factor which agreed greatly with and validated those estimated from the model. Having an accurate idea about the underreporting factor and the extent of spread is extremely helpful in terms of tracking the growth of the pandemic and determining policies. Repeated serological surveys to track the ever-evolving seroconversion scenario are often not viable options due to being expensive both in terms of cost, resources and time, the model estimates, updated regularly with new incoming data, provide an option to keep track of the underreporting factor and unreported cases and deaths.

The Delhi Serology Study is one of several COVID-19 serology studies that have been conducted across the world (for a summary of such studies, please refer to Table 3)^8, 17-28^. The seroprevalence found in the Delhi Serology Study (22.86%) is the highest among these studies but is similar to that found in New York City (22.70%), another large, densely populated area^26^. This indicates that Delhi is definitely on the higher side in terms of seroprevalence, even within worldwide epicenters and hotspots of COVID-19. Another serosurvey conducted in the city of Mumbai, India found strikingly different seroprevalences in slum (57.8%) vs non-slum areas (16%), the overall estimated seroprevalence also being staggeringly high (40.5%) compared to other studies across the world (Table 3)^19^.

**Table 3.** Summary of COVID-19 seroprevalence studies.

| Location | Study Design | Sample Size | Estimated Seroprevalence % (95% CI) | Reference |
| --- | --- | --- | --- | --- |
| Hubei and Guangdong Provinces<br>*China* | Cohort and location-specific surveys (Healthcare workers and their relatives, hospital outpatients, factory workers, hotel staff). | 6,919 (Hospital settings)<br>10,449 (Community settings) | 3.8 (2.6, 5.4) *Healthcare workers, Wuhan*<br>3.8 (2.2, 6.3) *Hotel staff members, Wuhan*<br>3.2 (1.6, 6.4) *Family members, Wuhan* | Xu et al. (2020)<br>*Nature Medicine*<br>doi: 10.1038/s41591-020-0949-6 |
| Essen<br>*Germany* | Prospective cross-sectional monocentric study recruiting healthcare workers from University Hospital Essen. | 316 | 1.6 | Korth et al. (2020)<br>*Journal of Clinical Virology*<br>doi: 10.1016/j.jcv.2020.104437 |
| India | Pilot survey in 83 districts across 21 states. | Unknown | 0.73 *overall*<br>1.09 *urban* | *The Indian Express* (2020)<br>url:<br>https://indianexpress.com/article/explained/delhi-serological-survey-shows-antibodies-in-23-participants-what-does-this-mean-6516512/ |
| Mumbai<br>*India* | Consent-based survey across three wards with high COVID-19 growth and proximity to hotspots. | 6,936 (Out of 8,800 invited) | 40.5 *overall*<br>57.8 *slum areas*<br>16.0 *non-slum areas* | *The Indian Express* (2020)<br>url:<br>https://indianexpress.com/article/explained/mumbai-serosurvey-what-it-shows-about-gender-differences-in-infection-mortality-and-herd-immunity-6529186/ |
| Guilan Province<br>*Iran* | Population-based cluster random sampling through phone call invitations. | 552 (196 households) | 0.22 (0.19, 0.26) *unadjusted*<br>0.33 (0.28, 0.39) *adjusted for imperfect testing*<br>0.21 (0.14, 0.29) *adjusted by population weights* | Shakiba et al. (2020)<br>*medRxiv*<br>doi: 10.1101/2020.04.26.20079244 |
| Kobe City<br>*Japan* | Cross-sectional study on hospital outpatients. | 1,000 | 3.3 (2.3, 4.6) | Doi et al. (2020)<br>*medRxiv*<br>doi: 10.1101/2020.04.26.20079822 |
| Spain | Two-stage random sampling of households stratified by province and municipality size. | 61,075 (Point-of-care test)<br>51,958 (Immunoassay)<br>(35,883 households) | 5.0 (4.7, 5.4) *point-of-care test*<br>4.6 (4.3, 5.0) *immunoassay* | Pollán et al. (2020)<br>*The Lancet*<br>doi: 10.1016/S0140-6736(20)31483-5 |
| Sweden (9 Regions) | Consecutive weekly region-specific surveys. | 1,200 (Per week). | 7.3 *Stockholm*<br>4.2 *Skåne*<br>3.7 *Västra Götaland* | Public Health Agency Sweden (2020)<br>url: https://www.folkhalsomyndigheten.se/nyheter-och-press/nyhetsarkiv/2020/maj/forsta-resultaten-fran-pagaende-undersokning-av-antikroppar-for-covid-19-virus/ |
| Geneva<br>*Switzerland* | Series of 5 consecutive weekly serosurveys among randomly selected participants from a previous population-representative survey, and their household members aged 5 years and older. | 2,766 (1,339 households; 341, 469, 577, 604 and 775 samples respectively in weeks 1-5.) | 4.8 (2.4, 8.0) *week 1*<br>8.5 (5.9, 11.4) *week 2*<br>10.9 (7.9, 14.4) *week 3*<br>6.6 (4.3, 9.4) *week 4*<br>10.8 (8.2, 13.9) *week 5* | Stringhini et al. (2020)<br>*The Lancet*<br>doi: 10.1016/S0140-6736(20)31304-0 |
| LA County, California<br>*USA* | Invited enrollment, based on demographic match and geographical proximity to the testing centers. | 863 (Out of 1952 invited) | 4.06 (2.84, 5.60) *unadjusted*<br>4.34 (2.76, 6.07) *adjusted for imperfect testing* | Sood et al. (2020)<br>*JAMA*<br>doi: 10.1001/jama.2020.8279 |
| New York State<br>*USA* | Convenience sampling of New Yorkers attending 99 grocery stores across 26 counties, containing 87.3% of the state's population, located all across the state. | 15,101 | 14.0 (13.3, 14.7) *overall*<br>22.7 (21.5, 24.0) *New York City* | Rosenberg et al. (2020)<br>*Annals of Epidemiology*<br>doi: 10.1016/j.annepidem.2020.06.004 |
| San Francisco Bay Area<br>*USA* | Cohort-based recruitment of non-COVID patients and blood donors. | 387 (Non-COVID patients)<br>1,000 (Blood donors) | 0.26 (0.00, 0.76) *non-COVID patients*<br>0.10 (0.00, 0.56) *blood donors* | Ng et al. (2020)<br>*medRxiv*<br>doi: 10.1101/2020.05.19.20107482 |
| Santa Clara County,<br>California<br>*USA* | Ad-based recruitment, matched on geographic location and demographics. | 3,330 | 1.5 (1.1, 2.0) *unadjusted*<br>1.2 (0.7, 1.8) *adjusted for imperfect testing*<br>2.8 (1.3, 4.7) *adjusted for county demographics* | Bendavid et al. (2020)<br>*medRxiv*<br>doi: 10.1101/2020.04.14.20062463 |

Extensive and long-drawn discussions have already taken place in relation to potential community transmission of COVID-19 in India. While even without a serological survey it is possible to comment on this based on other information available in terms of the number of tests and test positive rates, our results confirm that at least for Delhi, there is undoubtedly community transmission with regards to the classical definition of the term^29^. With more than 500,000 active cases in India and more than 10,000 active cases in Delhi as of July 28, many of the cases are potentially not being tracked to an identifiable source of infection. Along these lines, there have been debates about achieving herd immunity, and estimated range for the herd immunity threshold lies within 44-73% (based on worldwide estimated basic reproduction number of 1.8-3.8)^30-31^.

Both for Delhi and more so possibly for other parts of India, herd immunity will potentially take some time to be attained and is definitely not a panacea we can rely on. Even based on the IFR obtained without adjusting for potential death underreporting and trusting the reported death counts (Table 2), if 50% people in India, with a population of 1.38 billion, get infected (a concept that many proponents of herd immunity have suggested), it would imply an estimated **550**,**000** deaths, which skyrockets to an estimate of a staggering 5.5 million deaths if we believe the estimated underreporting factor from death from our proposed model.

There are several factors that we need to take into account about the Delhi Serological Study and consequently, these factors also shape the potential implications of our results. A large set of important information isn’t well-known or wasn’t publicly reported in the NCDC serology survey, such as the response and positivity rates stratified by age, sex, job type, district; sampling design; sensitivity or specificity of the customized assay – and so on. Releasing a single magic number without a complete report is definitely not the best practice for science and policy and is dissatisfying and potentially misleading. Further, we do not know if individuals with antibodies are protected from re-infection or how long this protection lasts^32^. We need to know more about the longevity of the antibody response and the levels of it needed to protect us from re-infections, and also about the contagiousness and potential clinical severity of a person with the antibody.

Even though the appearance and spread of COVID-19 has taken the entire world by a storm, a large number of examples from all across the world clearly depict that with extensive testing, contact tracing, use of masks, hand hygiene and social distancing we can change the narrative and course of this virus. For example, Delhi has seen tremendous success in turning the corner of the virus curve with the reproduction number at 0.74 (July 28) and the R staying below unity for about a month (Figures 5-6). Rapid and significant scientific advancements in both clinical and public health aspects of the disease have been made over the past few months^33^, and focused and cautious tracking of the pandemic with informed policy decisions are going to be as helpful as ever at this point. In that line, our analytical framework of integrating diagnostic testing imperfections in context of estimating unreported cases using the extended SEIR model and validating against seroprevalence estimates will hopefully prove to be useful for other case-studies.

**Figure 5.**
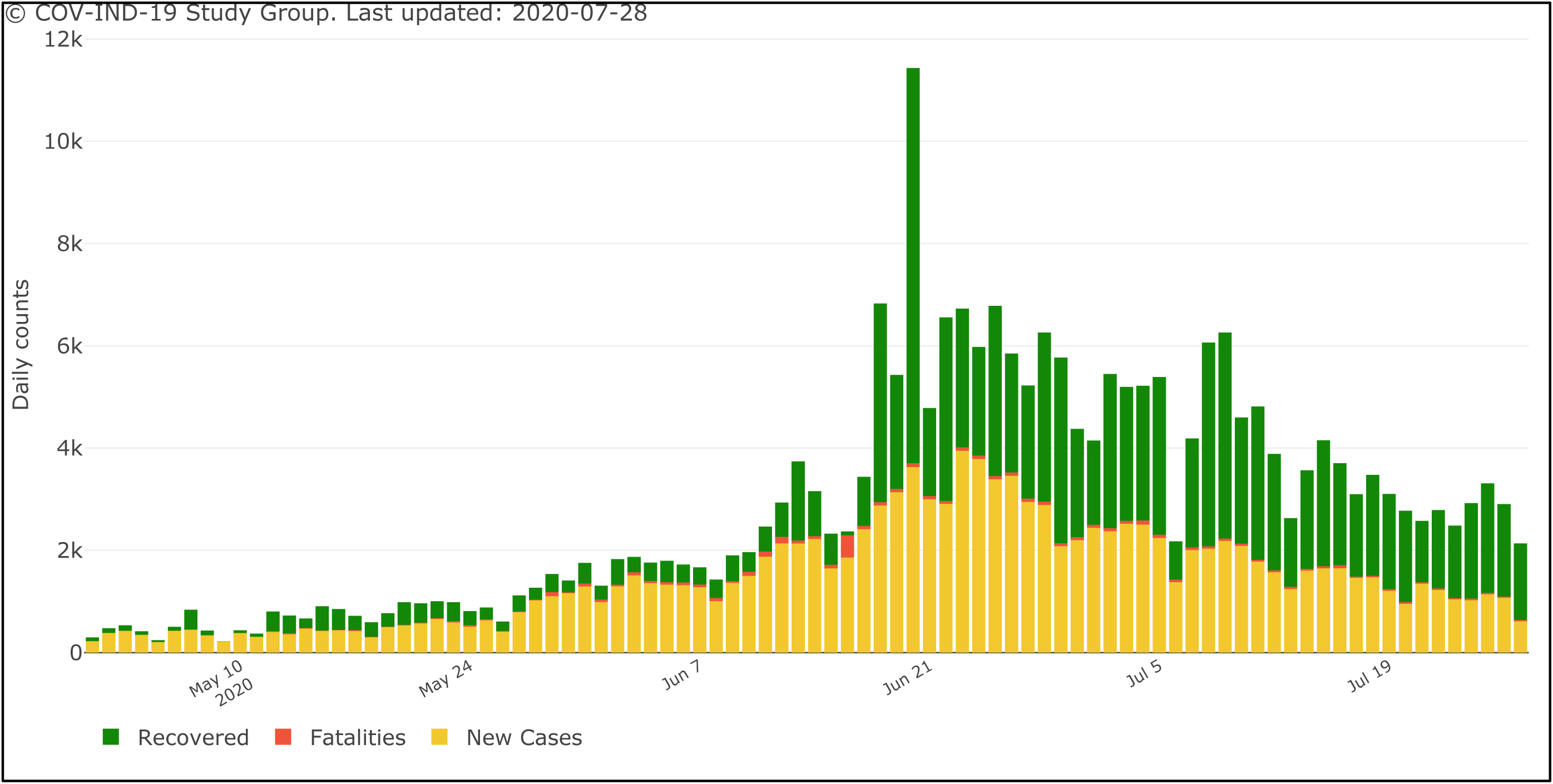
Observed daily COVID-19 case, recovery and fatality counts for Delhi during May 1 – July 28.

**Figure 6.**
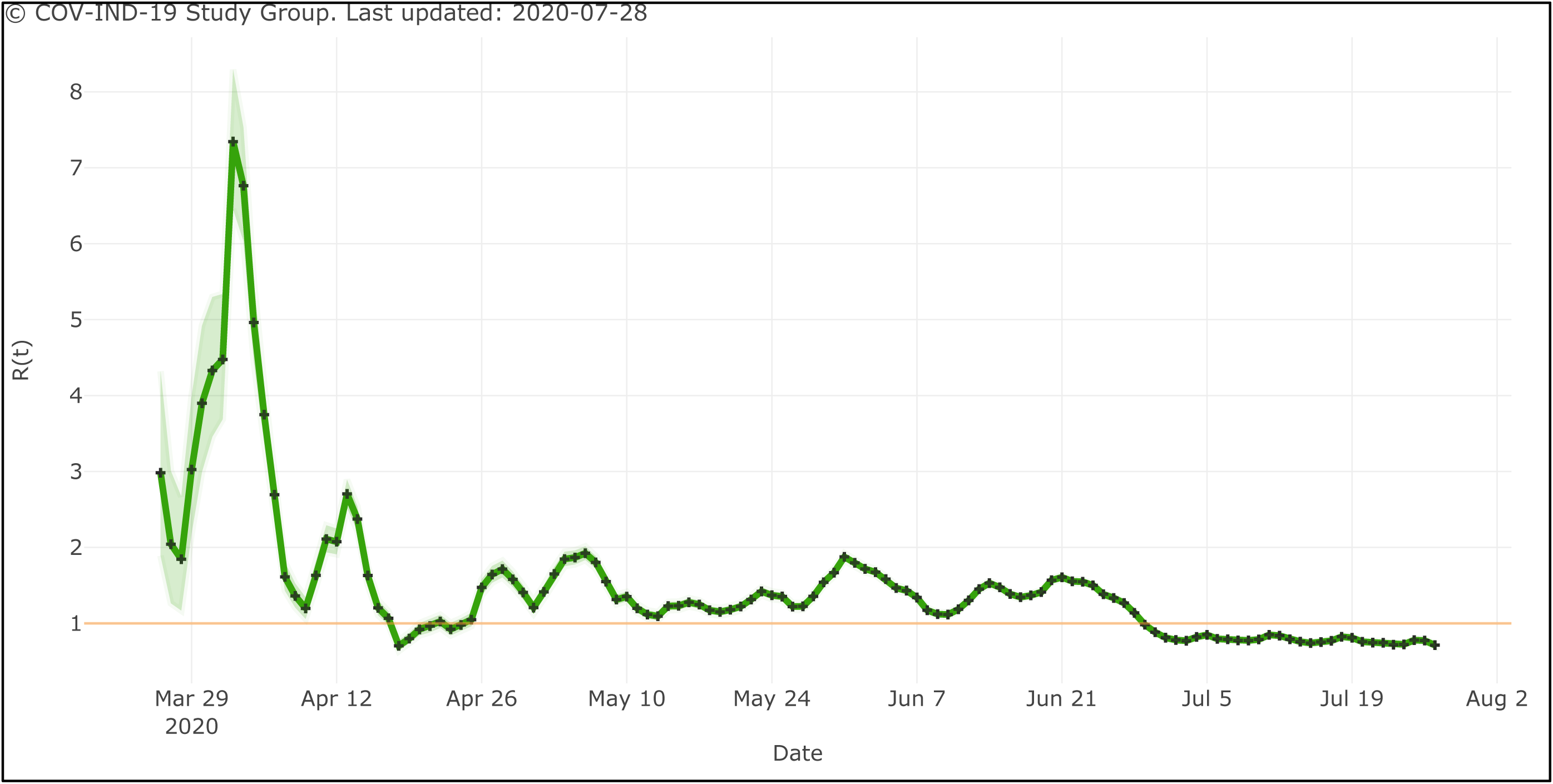
Estimated time-varying R (and 95% posterior credible interval) for COVID-19 in Delhi. Computations were performed using the R package EpiEstim.

## METHODS

### Extended SEIR Model Adjusted for Misclassification

We developed an extension of a standard SEIR model. In this model, the susceptible individuals (S) become exposed (E) when they are infected, but they have not started infecting the other people. After a latency period, exposed individuals are able to infect other susceptible individuals and are either untested (U) with probability *r* or tested (T) with probability 1 − *r*. Tested individuals enter either the false negative compartment (F) with probability *f* or the (true) positive compartment with probability 1 − *f*. Individuals who are in the untested and the false negative compartments are considered unreported COVID-19 cases and enter either the recovered unreported (RU) or death unreported (DU) compartments. Similarly, those who tested positive move to either a recovered reported (RR) or death reported (DR) compartment. Figure 7 represents the SEIR model schematic. The corresponding system of differential equations are presented below. The parameters and their values used are described in Table 4.

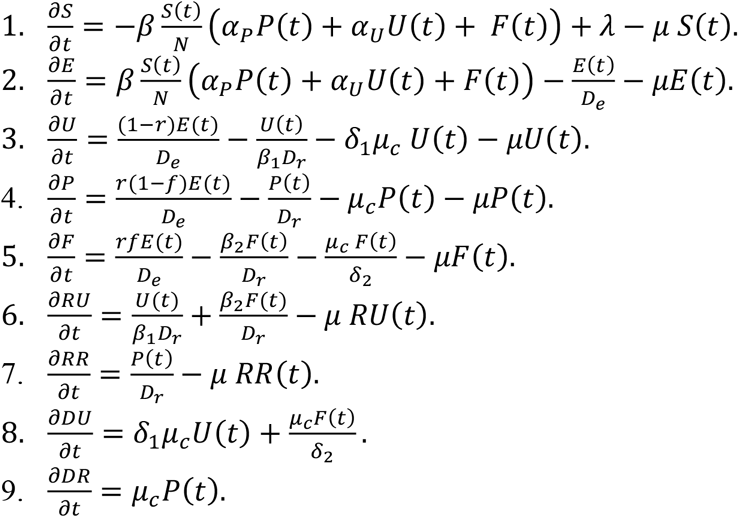

**Table 4.** Description of extended SEIR model parameters.

| Parameter | Value | Description |
| --- | --- | --- |
| $\beta$ | *Time-varying* | Rate of infectious transmission by infected, tested individuals with false negative results. |
| $\alpha_p$ | 0.5 | Ratio of rate of spread of infection by tested positive patients to that by false negatives. $\alpha_p < 1$ represents the scenario where individuals who test positive are infecting susceptible individuals at a lower rate than infected individuals with false negative test results. |
| $\alpha_u$ | 0.7 | Scaling factor for the rate of spread of infection by untested individuals. $\alpha_u$ is assumed to be $< 1$ as U mostly consists of asymptomatic or mildly symptomatic cases who are known to spread the disease at a much lower rate than those with higher levels of symptoms. |
| $D_e$ | 5.2 | Incubation period (in days). |
| $D_r$ | 17.8 | Means number of days until recovery for infected individuals. |
| $D_t$ | 0 | Mean number of days for the test result to come after a person is tested. Under the assumption of instantaneous test results, this is taken to be zero. |
| $\mu_c$ | 0.0562 | Death rate attributable to COVID-19 which is equivalent to inverse of the average number of days for death starting from the onset of disease times the probability of death of an infected individual. |
| $\lambda, \mu$ | $3.95 \times 10^{-5}$ | Natural birth and death rates (assumed to be equal). |
| $r$ | *Time-varying* | Probability of being tested for infectious individuals. |
| $f$ | 0.3, 0.15, 0 | Probability of a false negative RT-PCR diagnostic test result. |
| $\beta_1, \frac{1}{\beta_2}$ | 0.6 ($\beta_1$)<br>0.7 ($\beta_2$) | Scaling factors for rate of recovery for undetected and false negative individuals respectively. Both $\beta_1$ and $\beta_2$ are assumed to be less than 1. It is assumed that the recovery rate is slower than the detected ones for the False Negative ones because they are not getting any hospital treatments. The condition of Untested individuals is not so severe as they consist of mostly asymptomatic people. So, they are assumed to recover faster than the Current Positive Ones. |
| $\delta_1, \frac{1}{\delta_2}$ | 0.3 ($\delta_1$)<br>0.7 ($\delta_2$) | Scaling factors for death rate for undetected and false negative individuals respectively. Both $\delta_1$ and $\delta_2$ are assumed to be less than 1. Same as before, the death rate for False Negative ones are assumed to be higher than the Current detected Positive as they are not receiving proper treatment. While, for the Untested ones, the death rate is taken to be lesser because they are mostly asymptomatic. So, their probability of dying is much less. |

**Figure 7.**
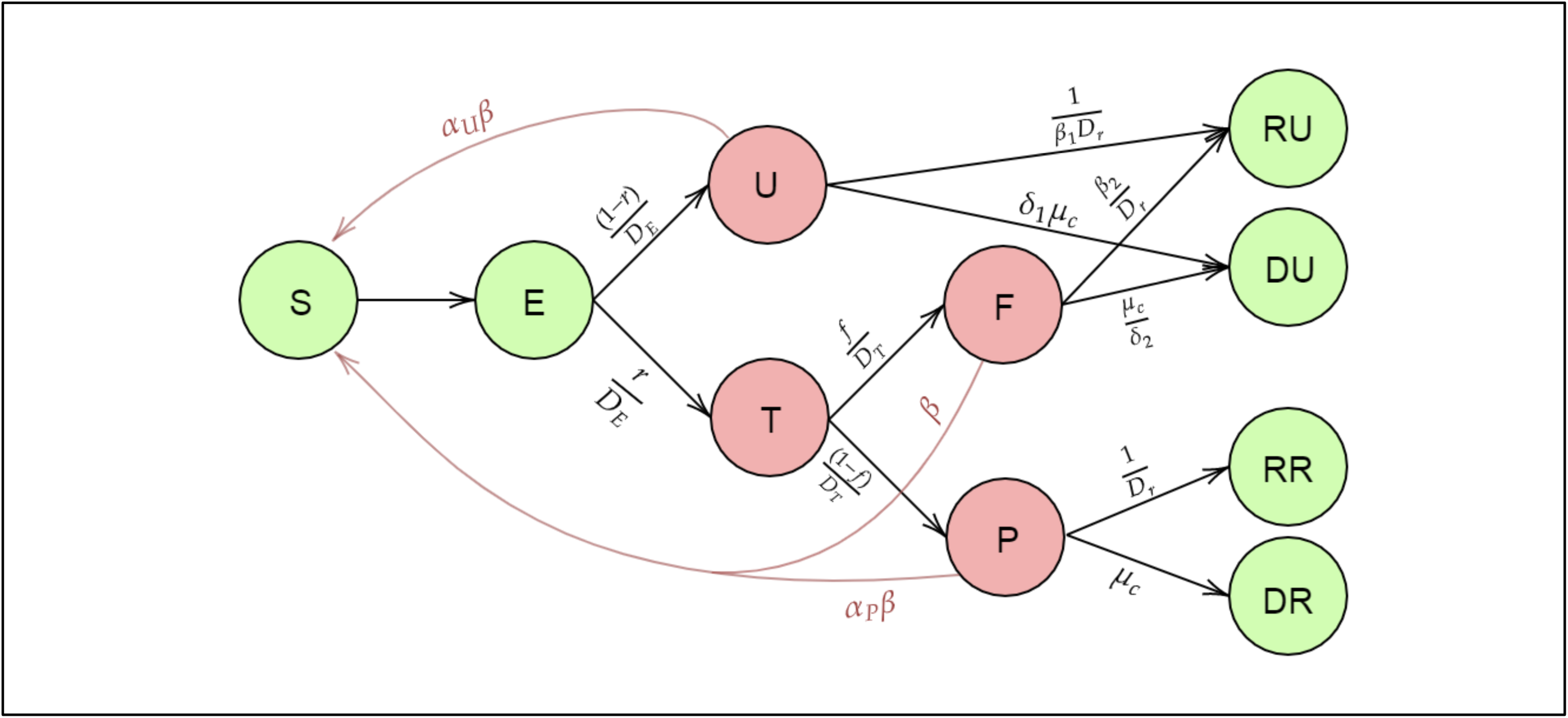
Diagram of model compartments and transmissions for the extended SEIR model.

Here, *X* (*t*) denotes the number of individuals in the compartment of interest *X* at time *t*. In this setting, both *β* and *r* are time-varying parameters which are estimated using the Metropolis-Hastings MCMC method^34^. To estimate the parameters, we first need to be able to solve the differential equations, which is difficult to perform in this continuous-time setting. It is also worth noting that we do not require the values of the variables for each time point, we only need their values at discrete time steps, i.e., for each day. Thus, we approximate the above set of differential equations by a set of recurrence relations. For any compartment *X*, the instantaneous rate of change with respect to time A (given by 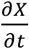) is approximated by the difference between the counts of that compartment on the (*t* + 1)^*th*^ day and the *t*^*th*^ day, that is *X* (*t* + 1) − *X* (*t*). Starting with an initial value for each of the compartments on the Day 1 and using the discrete-time recurrence relations, we can then obtain the solutions of our interest. Some examples of these discrete-time recurrence relations are presented below.

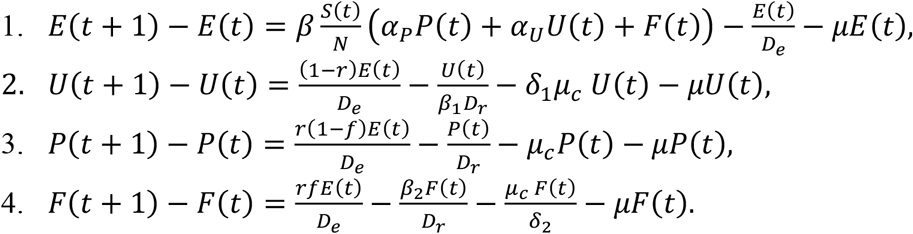

The rest of the differential equations can be similarly approximated by a discrete-time recurrence relation. These parameters are estimated using training data from Delhi from March 15 to June 30. The training data to divided into 7 periods, in accordance with the lockdown and unlock procedures employed by the government of India, as described in Table 5. Using these, we performed our predictions for the dates ranging from June 1 through July 26.

**Table 5.**
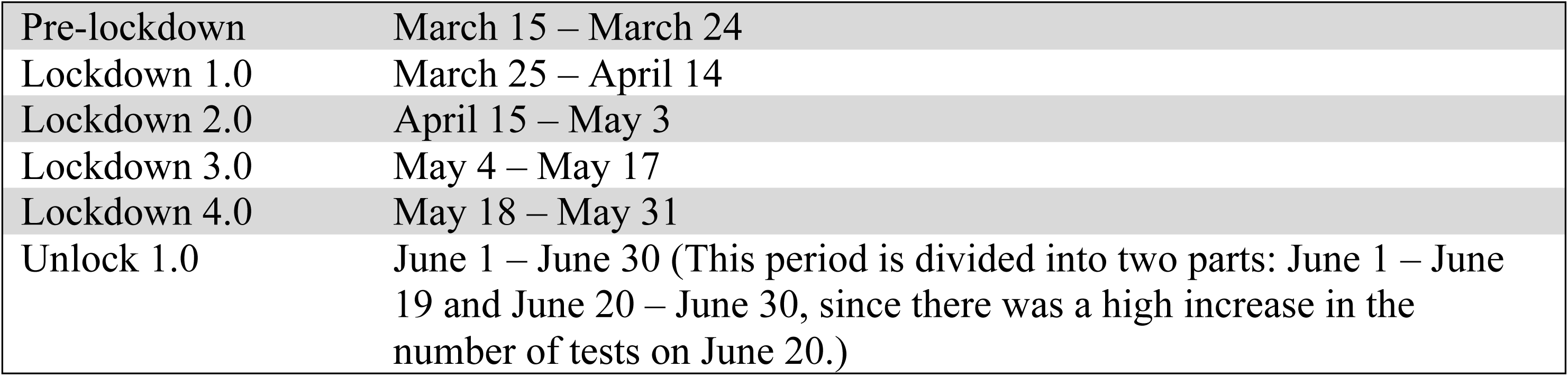
Training data periods according to interventions.

|  |  |
| --- | --- |
| Pre-lockdown | March 15 – March 24 |
| Lockdown 1.0 | March 25 – April 14 |
| Lockdown 2.0 | April 15 – May 3 |
| Lockdown 3.0 | May 4 – May 17 |
| Lockdown 4.0 | May 18 – May 31 |
| Unlock 1.0 | June 1 – June 30 (This period is divided into two parts: June 1 – June 19 and June 20 – June 30, since there was a high increase in the number of tests on June 20.) |

### Naïve Corrections to Reported Test Results using Known Misclassification Rates

Let us set up a few notations first. Let N = population size, X = number of true cases in the population (hence N – X = number of non-cases in the population), T = number of people tested, S = number of true cases tested (hence T – S = number of non-cases tested, X – S = number of true cases not tested, N – X – T + S = number of non-cases not tested), P = number of positive tests (also, therefore, cumulative number of reported cases, hence T – P = number of negative tests). Note that X and S are the only two unknowns in this setting. Also, let us assume that the sensitivity of the test of interest is *α* and the specificity of the same is *β*. With that, we can set up the following equation, because there are two ways a test can be positive, as can be seen in Figure 8.

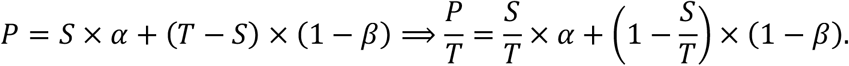

**Figure 8.**
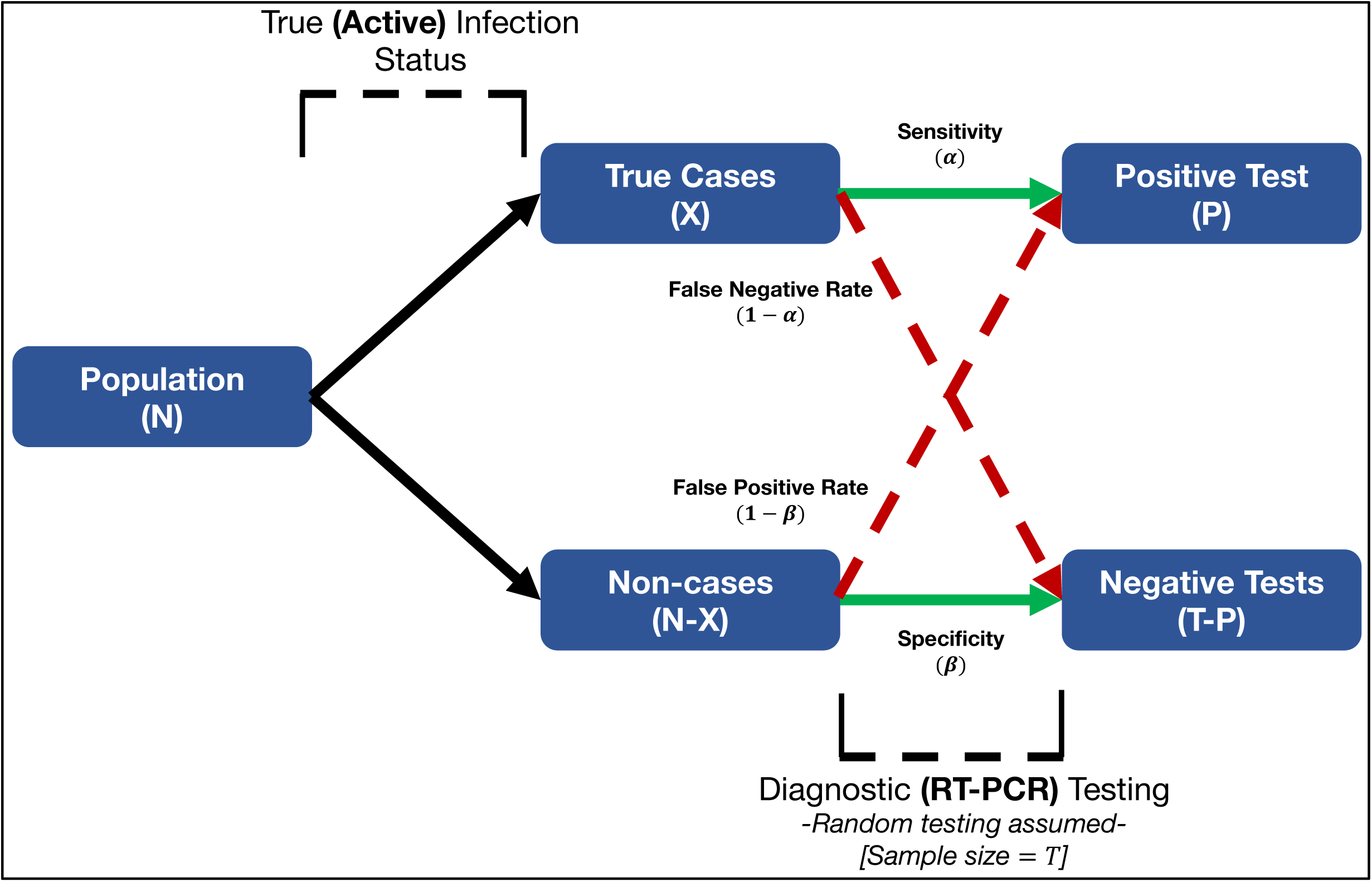
Diagram of testing decisions. Dark lines indicate the break-up of the population in terms of true infection status; green and red lines indicate (correct and incorrect, respectively) decisions based on testing procedure. Here, we have referred to the diagnostic test, and hence, the active infection status. Similar framework applies to the antibody test and the past infection status.

Adjusting the terms, we get the following expression for *S*.

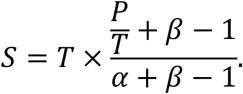

Assuming that the proportion of cases among those tested stays the same as the original population (random and hence homogenous testing), we can replace *S* by 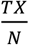, which will lead to the following updated equation.

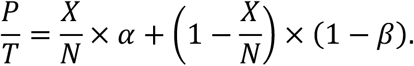

Solving this, we get the following expression for *X*.

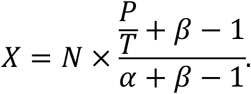

Thus, these two expressions give us, for a given set of *α* and *β* regarding a test, the corrected number of reported cases (*S*), and also the estimated number of true (reported + unreported) cases (*X*). For the computation of *S*, we use 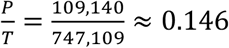, the test positive rate of the RT-PCR tests in Delhi as of July 10^2^. For the computation of *X*, we use 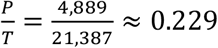, the positive rate reported by the Delhi serological survey^7-9^. Once we get these two estimates, we can compute the adjusted underreporting factor as 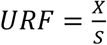. Also, assuming that *D* denotes the cumulative number of deaths till a date of interest, we can compute the corrected versions of case fatality rate and infection fatality rate as 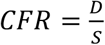 and 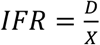, respectively. Further, if we want to adjust for a potential scenario where for every M deaths due to COVID-19, we observe 1 death (M-fold underreporting for deaths), we can update these estimates as 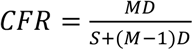 and 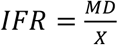. We computed our adjusted CFR and IFR estimates for *M* = 10. Based on the data till July 10 from Delhi, we use *D* = 3,300^2^. We also used a population size of *N*= 1.98 × 10^7^.

A critical question here is the choice of *α* and *β* for the two tests to ensure our computations reflect adjustments made based on sensible and realistic scenarios. Based on previously reported sensitivity and specificity levels for the diagnostic test^13^, we used the combinations *α* = *β* = 1 (*perfect test*), *α* = 0.85 *and β* = 0.99, *and α* = 0.7 *and β* = 0.99. The serological assay used by NCDC is a customized assay, for which we could not find any officially reported numbers. Hence, we referred to existing literature on standard serological assays^14^ and publicly available discussions on the Delhi serosurvey^15^, and decided to use the combinations of *α* = *β* = 1 (*perfect test*), *α* = 0.976 *and β* = 0.993, *and α* = 0.92 *and β* = 0.97.

## Data Availability

All our computational codes are available at covind19.org.

http://covind19.org/

## CODE AVAILABILITY

All our computational codes are available at covind19.org.

## ACKNOWLEDGEMENTS

The authors would like to thank the Center for Precision Health Data Sciences at the University of Michigan School of Public Health, The University of Michigan Rogel Cancer Center and the Michigan Institute of Data Science for internal funding that supported this research. The authors are grateful to Professors Eric Fearon, Aubree Gordon and Parikshit Ghosh for useful conversations that helped formulating the ideas in this manuscript.

## AUTHOR CONTRIBUTIONS

Rupam Bhattacharyya prepared the initial draft and carried out the naïve misclassification correction to case counts and took leadership of composing the final draft. Ritwik Bhaduri, Ritoban Kundu and Bhramar Mukherjee developed the extended SEIR model with misclassification. Ritwik Bhaduri and Ritoban Kundu implemented the extended SEIR model. Maxwell Salvatore carried out extensive literature search and visualization and participated in writing of the manuscript. Bhramar Mukherjee conceptualized the project and oversaw the research.

## MATERIALS AND CORRESPONDENCE

All correspondences should be directed to Bhramar Mukherjee. All data and code are available at covind19.org.

